# Associations of HIV and prevalent type 2 diabetes mellitus in the context of obesity in South Africa

**DOI:** 10.1101/2024.03.10.24304033

**Authors:** IM Magodoro, AC Castle, N Tshuma, JH Goedecke, R Sewpaul, J Manasa, J Manne-Goehler, NAB Ntusi, MJ Nyirenda, MJ Siedner

## Abstract

It is unclear how rising obesity among people with HIV (PWH) in sub-Saharan Africa (SSA) impacts their risk of type 2 diabetes mellitus (diabetes). Using a South African national cross-sectional sample of adult PWH and their peers without HIV (PWOH), we examined the associations between HIV and prevalent diabetes across the spectrum of body mass index (BMI), waist circumference (WC) and waist-to-height ratio (WtHR). Analyses were sex stratified, and adjusted for age, sociodemographic and behavioral factors. The prevalence of diabetes among males was similar between PWH and PWOH, overall and at all levels of adiposity. In contrast, overall diabetes prevalence was higher among female PWOH than female PWH. However, there were differences according to adiposity such that, compared to female PWOH, relative diabetes prevalence in female PWH was reduced with obesity but accentuated with leanness. These differences in the relationship between adiposity and diabetes by HIV serostatus call for better mechanistic understanding of sex-specific adipose tissue biology in HIV in South Africa, and possibly in other HIV endemic settings in SSA.

## Introduction

The anticipated pandemic of HIV-related cardiometabolic disorders in sub-Saharan Africa (SSA) has not yet become evident, 20 years into the era of widely accessible antiretroviral treatment (ART).^1, 2^ In high-income countries (HICs), treated HIV is a recognized risk factor for numerous cardiovascular and metabolic disorders, such as heart failure, stroke, metabolic syndrome, obesity, hypertension, and type 2 diabetes mellitus (diabetes), among others.^3, 4, 6^ People with HIV infection (PWH) in HICs, for example, have greater risk of both prevalent^7, 8, 9^ and incident^10, 11, 12^ diabetes than comparable persons without HIV (PWOH). It was expected that a similar upsurge will occur in SSA as ART uptake increases.^13, 14,^

However in SSA, PWH who are on ART, including older-generation thymidine analogues and protease inhibitors, appear to have comparable^15, 16^ if not better^17, 18, 19^ glucose metabolism profiles than the general population. While this may be attributed to insufficient research in SSA, especially the dearth of longitudinal studies with incidence measures^20^, physiological adaptations in PWH in SSA that potentially protect against cardiometabolic disease, including diabetes, have not been considered.

In the recent landmark REPREIVE trial (2023), major adverse cardiovascular events were noted to be higher in HICs (10.7 per 1,000 person-years) than in SSA (1.8 events per 1,000 person-years).^21^ Relatedly, Brenan *et. al.,* (2023) using country-wide data from 373,889 patients tested for diabetes through South Africa’s National Health Laboratory Service, found PWH were less likely to have diabetes (adjusted relative risk: 0.73; 95% CI: 0.71–0.74) than PWH.^19^ If indeed ART-experienced PWH in SSA, unlike their counterparts in HICs, are spared from excess cardiometabolic disease risk, particularly diabetes, it is imperative to understand the underlying mechanisms. Given the rising diabetes rates in the general population^22, 23^ such insights may inform novel therapeutic and risk stratification approaches in SSA and HICs.

Obesity is integral to diabetes pathogenesis, with over 80% of individuals with diabetes in the general population in HICs living with overweight or obesity (body mass index (BMI) ≥25 kg/m^2^).^24, 25^ Among the metabolic consequences of obesity are adipose tissue inflammation, disordered adipokine signaling, mitochondrial dysfunction, dyslipidemia including increased circulating free fatty acids (FFAs), and ectopic fat deposition.^25, 26^ These in turn drive β-cell dysfunction as well as insulin resistance^26, 27^, the two gateways to impaired glucose tolerance and diabetes.^28^ On the other hand, HIV itself and the side effects of ART, particularly for older generations of therapy, may directly and indirectly cause similar metabolic derangements.^29, 30^ with more modern regimens, weight gain following ART initiation often surges beyond “return to health” into deleterious categories of excess adiposity.^31, 32^ This includes central obesity, with or without lipodystrophy, both significant determinants of prevalent and new-onset diabetes.^31^ It is the nexus of these factors, i.e., HIV, ART metabolic side effects, excess weight gain and an obesogenic environment, that is believed to drive excess diabetes risk in persons PWH in HICs^10, 11, 12^ whereas its occurrence in SSA^17, 18, 33^, is yet to be mirrored in an excess burden of diabetes in the population of PWH.

Understanding the interplay of obesity, HIV, and ART’s metabolic effects in SSA is essential to elucidate the unique factors influencing diabetes prevalence in this region. We used data from a national survey in South Africa to examine whether HIV is associated with diabetes prevalence, and if such associations vary with obesity.

## Methods

We followed the guidelines of the Strengthening the Reporting of Observational Studies in Epidemiology (STROBE) in the conduct and reporting of our analyses.^34^

### Study population and analytic sample

The present study is a secondary analysis of the 2016 South Africa Demographic Survey (SADHS), which is fully described elsewhere.^35^ The 2016 SADHS is a cross-sectional survey of the health status, and its determinants, of noninstitutionalized South Africans. Sampling followed a stratified two-stage design with primary sampling units drawn from urban, rural, and farming areas. The division of South Africa into urban, rural and farming areas accounts for its historical race-based spatial socio-economic development. Data were collected between June and November 2016 using questionnaires, physical examination, and laboratory testing of blood. Only participants aged at least 20 years with complete information on HIV testing, anthropometry, and HbA1c were eligible for inclusion in the present analysis.

### Study Measures

#### Glucose metabolism

Finger prick blood was tested for glycated hemoglobin (HbA1c) using the Roche Tina-quant® II immunoturbidimetric assay on a blood chemistry analyzer (Hitachi 912 Analyzer, Hitachi, Tokyo, Japan). In addition to HbA1c as a continuous study endpoint, we defined diabetes as any current use of clinician-prescribed oral hypoglycemic medicines and/or insulin and/or HbA1c ≥6.5%.^36^ A second definition of diabetes, HbA1c ≥6.5% in a subsample (age ≥40 years old) without self-reported current use of oral hypoglycemic medicines or insulin, was used for the sensitivity analysis.

#### HIV serostatus

HIV serostatus was ascertained from dry blood spots using an HIV antibody testing algorithm with three different enzyme-linked immunosorbent assays (ELISA). Concordant negative results on the ELISA 1 (Genscreen HIV 1/2 Combi Assay, Bio-Rad, France) and ELISA 2 (E411 Cobas HIV 1/2 Combi Assay, Roche, Switzerland) were recorded as negative. Discordant ELISA 1 and ELISA 2 results were repeated, and if still discordant, were classified as indeterminant. Concordant positive results on the ELISA 1 and ELISA 2 were classified as positive if a third confirmatory test (Geenius™ HIV1/2 confirmatory rapid test, Bio-Rad, France) was positive, and inconclusive if the confirmatory test was negative or indeterminate.^35^

#### Body mass index and waist circumference

Weight and height were measured using portable digital scales (Seca 878 dr, Seca, Hamburg, Germany) and stadiometers (Seca 213 dr, Seca, Hamburg, Germany), respectively, while waist circumference (WC) was measured using non-stretch tape (Seca 203 dr, Seca, Hamburg, Germany) at the level mid-way between the lowest rib margin and the iliac crest with the subject standing erect and at the end of gentle expiration. Body mass index (BMI) estimated from weight (in kilograms)/height (in meters)^2^ was classified as underweight (<18.5kg/m^2^), normal (18.5–24.9kg/m^2^), overweight (25.0–29.9kg/m^2^), and obese (≥30.0kg/m^2^). Waist circumference was categorized as elevated if ≥94cm for males and ≥80cm for females, or normal if <94cm for males and <80cm for females.^37^ Lastly, we defined waist-to-height ratio (WtHR) (waist circumference/height) ≥0.5 as elevated.^38^

#### Sociodemographic and other covariates

Sex was self-reported as male or female. Participants described their race/ethnicity as any one of Black, White, Colored (for mixed race), or Indian/Asian, while their place of residence was classified as either urban or rural. Residence in farming areas was considered rural. Socioeconomic position was indicated by the household wealth index. The latter is a composite measure of a household’s cumulative living standard calculated using principal components analysis^39^ and based on ownership of selected household assets such as television, radio, refrigerator, and vehicle; materials used for housing construction; and access to sanitation facilities and clean water. Cigarette use daily or on some days of the week was categorized as current smoking versus no cigarette use which was categorized as nonsmoking. Current drinkers were defined as those reporting alcohol consumption in the last 12 months; whereas non-drinkers were those reporting no alcohol consumption in the last 12 months. We reported blood pressure (BP) as the average of the last two of three automated (Omron 1300, Omron Healthcare, Bannockburn, IL, USA) resting state BP readings, and hypertension as any of systolic BP ≥140 mmHg, diastolic BP ≥90 mmHg or self-reported use of prescribed antihypertensive medication. Comorbid cardiovascular disease (CVD), chronic respiratory disease and past tuberculosis (TB) were also noted. These were defined based on self-reported clinician-led diagnoses of any of heart failure, stroke, or coronary artery disease for CVD; asthma, chronic bronchitis or chronic obstructive pulmonary disease chronic respiratory disease for chronic respiratory disease; and any completed drug treatment for TB.

### Ethics Review

The South African Medical Research Council’s Research Ethics Committee and the institutional review board of ICF approved the 2016 SADHS and all its data collection procedures, and written informed consent was obtained prior to participation. As a secondary data analysis, our study did not require any ethics review or approval. These data are publicly available in a de-identified form.

### Statistical Analysis

We performed a complete case analysis given negligible missingness (<2%) in HIV serostatus, HbA1c, sex, BMI, WtHR and WC data. Our outcome of interest was glucose metabolism which we modelled as prevalent diabetes (dichotomous endpoint) and mean HbA1c (continuous endpoint). Because our primary goal was to understand the complex relationships between diabetes, HbA1c and anthropometric indices between PWH and PWOH, and not to estimate population level parameters, we did not apply inverse probability sampling weights to our analysis. We described participants’ characteristics according to HIV serostatus. Raincloud plots were used to visualize the distribution of HIV and sex stratified HbA1c, with mean differences examined by linear regression. Next, sex-stratified multivariable fractional polynomial (MFP) generalized linear models were used to explore HIV-specific associations between HbA1c and each of (continuous) BMI, WtHR and WC. MFP modeling provides flexible parameterization to reveal non-linear associations^40^, and was thus suited to our hypothesis of complex relationships between HbA1c and obesity indices.

We also modeled the association between diabetes and (separately) categorical BMI, WtHR and WC using similar MFP generalized linear models stratified sex and including HIV as a predictor. Diabetes prevalence for each obesity category was derived from postestimation margins from these models. The corresponding PWH versus PWOH prevalence differences (PD) and prevalence ratios (PR) of diabetes across obesity categories were estimated from the postestimation margins and multiple linear combinations (for PD) or non-linear combinations (for PR) using Stata 18’s *mlincom* and *nlcom* commands, respectively.

#### Sensitivity Analyses

Sensitivity analyses re-examined the sex-stratified association between HIV and diabetes, and reported overall adjusted prevalence differences and prevalence ratios. The first approach used generalized linear models after propensity score (PS) matching on the same covariates as were used in primary analyses. The second approach used MFP generalized linear models adjusting for the same covariates as were used in the primary analyses. However, this time diabetes was defined as HbA1c ≥6.5% and analysis was restricted to a subsample of adults ≥40 years old not using oral hypoglycemic medicines or insulin. The PS matching approach assessed how sensitive our findings were to differences in data analysis methods and their attendant assumptions. With a higher cut-off age (≥40 years), the prevalence of undiagnosed type 1 diabetes and thus subsequent confounding is likely to be low, as is excluding treated diabetes which may indicate differential access to healthcare.

Analyses were conducted using R, version 3.6.3 (R Foundation for Statistical Computing, Vienna, Austria), and Stata version 18.0 (StataCorp, College Station, TX, USA). All probability values were 2-sided, with p-values <0.05 considered indicative of statistical significance.

## Results

### Sample characteristics

The analytic sample consisted of 5,635 total participants, of whom 4,381 (77.7%) were PWOH and 1,254 (22.3%) were PWH (**Table 1**). Mean age (95%CI) was 42 years (27, 59) for PWOH and 38 years (31, 47) for PWH. PLWH were more frequently females (73.7% vs. 60.9%) and of Black race (97.8% vs. 84.4%) than PWOH. While rates of urban/rural residence were comparable, PWH came from poorer households than PWOH. Smoking and alcohol consumption were similar between the two groups, as were the anthropometric measures. Overall, over half of participants had overweight/obesity (58.5%) according to BMI, and central obesity (52.6%) according to waist circumference. Comorbidities were also otherwise equally prevalent between the two groups except for diabetes (which is discussed below), treated TB, which was nearly threefold more common in PWH (14.7%) than PWOH (4.8%), and hypertension which was less common among PWH (46.6%) than PWOH (53.3%).

**Table 1.** Sample characteristics by HIV serostatus among adults aged ≥20 years old: unweighted South Africa Demographic Health Survey, 2016. Values are mean (SD) or ^α^median (25^th^, 75^th^ percentile) or number (percent). Elevated waist circumference if ≥94cm for males and ≥80cm for females. Elevated waist-to-height ratio if ≥0.5. PWH = people with HIV; PWOH = people without HIV.

| Characteristic | Total | HIV Serostatus |  |
| --- | --- | --- | --- |
|  |  | PWOH | PWH |
| Number | 5,635 | 4,381 | 1,254 |
| ***Sociodemographic*** |  |  |  |
| Male sex | 2,041 (36.2%) | 1,711 (39.1%) | 330 (26.3%) |
| Age (years) | 41.0 (28.0, 56.0) | 42.0 (27.0, 59.0) | 38.0 (31.0, 47.0) |
| Self-reported race |  |  |  |
| Black | 4,939 (87.7%) | 3,712 (84.8%) | 1,227 (97.8%) |
| White | 222 (3.9%) | 221 (5.0%) | 1 (0.1%) |
| Colored | 423 (7.5%) | 398 (9.1%) | 25 (2.0%) |
| Asian/Indian | 48 (0.9%) | 47 (1.1%) | 1 (0.1%) |
| Urban residence | 2,906 (51.6%) | 2,272 (51.9%) | 634 (50.6%) |
| Rural | 2,729 (48.4%) | 2,109 (48.1%) | 620 (49.4%) |
| Household asset index | 1.4 (-5.7, 6.9) | 2.1 (-4.8, 7.5) | -0.7 (-8.6, 4.7) |
| ***Behavioral*** |  |  |  |
| Current smokers | 1,032 (18.3%) | 843 (19.2%) | 189 (15.1%) |
| Non-smokers | 4,603 (81.7%) | 3,538 (80.8%) | 1,065 (84.9%) |
| Current drinkers | 1,802 (32.0%) | 1,450 (33.1%) | 352 (28.1%) |
| Non-drinkers | 3,833 (68.0%) | 2,931 (66.9%) | 902 (71.9%) |
| ***Anthropometric*** |  |  |  |
| Overweight/obesity | 3,243 (58.5%) | 2,544 (59.0%) | 699 (56.6%) |
| Overweight | 1,457 (26.3%) | 1,117 (25.9%) | 340 (27.6%) |
| Obesity | 1,786 (32.2%) | 1,427 (33.1%) | 359 (29.1%) |
| Elevated waist circumference | 2,939 (53.3%) | 2,286 (53.5%) | 653 (52.6%) |
| Elevated waist-to-height ratio | 3,371 (61.2%) | 2,642 (61.9%) | 735 (59.3%) |
| BMI (kg/m<sup>2</sup>) | 27.2 (6.4) | 27.8 (7.2) | 27.1 (6.7) |
| Waist circumference (cm) | 87.5 (14.8) | 87.7 (14.7) | 85.4 (13.0) |
| Waist-to-height ratio | 0.54 (0.10) | 0.54 (0.10) | 0.53 (0.09) |
| ***Cardiometabolic*** |  |  |  |
| Prevalent diabetes mellitus | 805 (14.4%) | 693 (15.9%) | 112 (9.0%) |
| HbA1c $\geq 6.5\%$ | 769 (13.6%) | 662 (15.1%) | 107 (8.5%) |
| HbA1c (%) | 6.2 (6.2, 6.3) | 6.3 (6.2, 6.3) | 6.1 (6.0, 6.1) |
| Prevalent hypertension | 2801 (51.8%) | 2,245 (53.3%) | 556 (46.6%) |
| Systolic BP (mmHg) | 132.8 (23.0) | 134.4 (23.7) | 127.5 (20.8) |
| Diastolic BP (mmHg) | 86.0 (12.7) | 86.0 (12.8) | 86.0 (12.4) |
| ***Self-reported comorbidities*** |  |  |  |
| Past drug treated tuberculosis | 392 (7.0%) | 208 (4.8%) | 184 (14.7%) |
| Cardiovascular disease | 227 (4.0%) | 183 (4.2%) | 44 (3.5%) |
| Chronic respiratory disease | 230 (4.1%) | 192 (4.4%) | 38 (3.0%) |

Values are mean (SD) or <sup>a</sup>median (25<sup>th</sup>, 75<sup>th</sup> percentile) or number (percent).

### Association between HbA1c, HIV and obesity indices

Stratified by HIV serostatus only, PWH had lower mean (95%CI) HbA1c [6.1% (6.0, 6.1)] than PWOH [6.3% (6.2, 6.3); p<0.001] (**Table 1**). In sex-specific comparisons, however, differences in mean HbA1c were seen only between female PWH [6.0% (5.9, 6.1)] and female PWOH [6.4% (6.3, 6.5); p<0.001], but not male PWH [6.1% (5.9, 6.2)] and male PWOH [6.1% (5.9, 6.2); p=0.234] (**Figure 1)**. **Supplementary Figures 2a-f** and **Figures 2a-f** depict, respectively, unadjusted and adjusted sex stratified HIV-specific associations between HbA1c and each of BMI, WtHR and WC. Among men, unadjusted and adjusted mean HbA1c had a largely linear relation to each of BMI, WtHR and WC with no differences by HIV serostatus.

**Figure 1.**
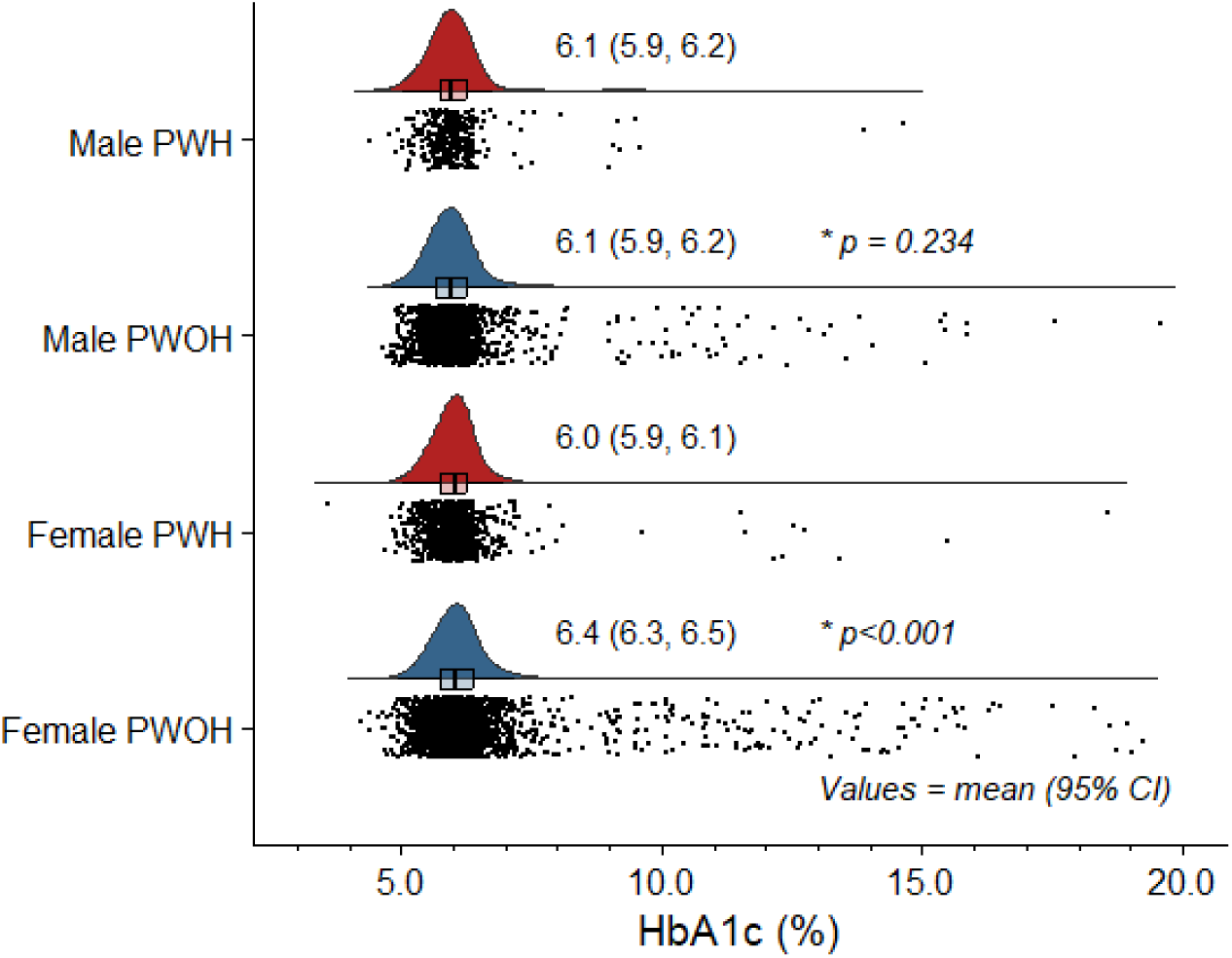
Distribution of glycated hemoglobin (HbA1c) according to HIV serostatus and sex among adults aged ≥20 years old: unweighted South Africa Demographic Health Survey, 2016. * P values for sex-specific persons PWH versus persons PWOH unadjusted comparisons of mean HbA1c level. Abbreviations as elsewhere defined.

**Figure 2.**
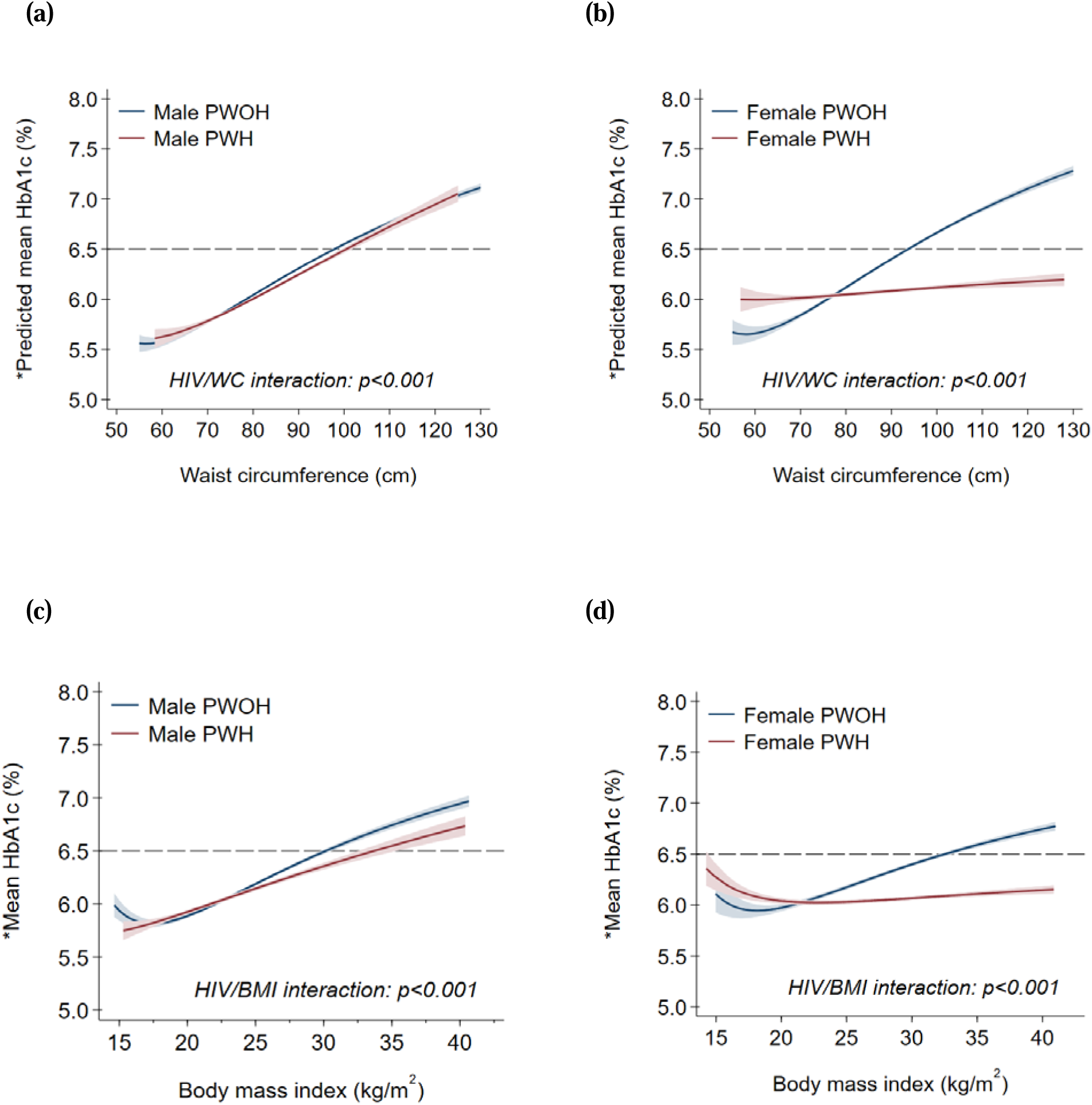
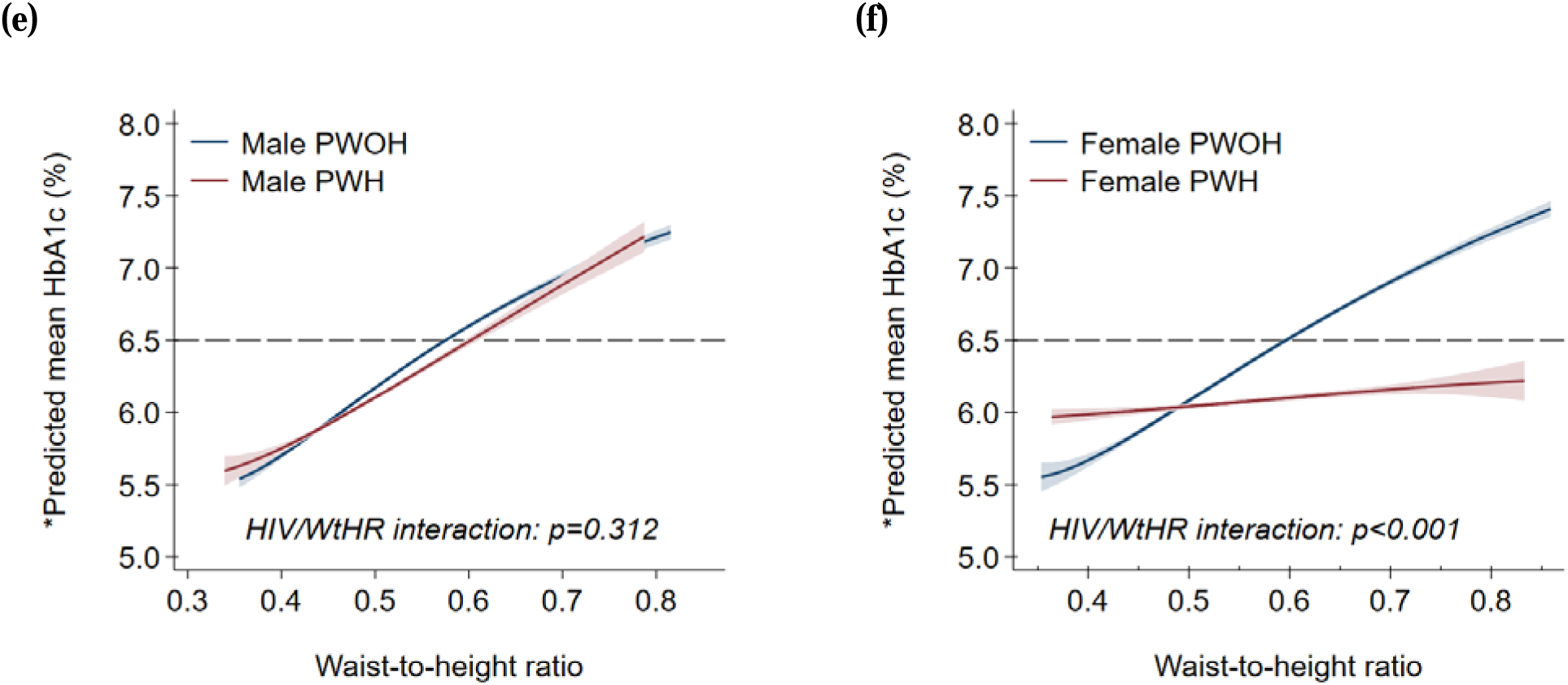
Predicted mean glycated (HbA1c) according to waist circumference, body mass index and waist-to-height ratio by HIV serostatus and sex among adults aged ≥20 years old: unweighted South Africa Demographic Health Survey, 2016. * Predicted from multivariable fractional polynomial models adjusted for age, sex, self-reported race, area of residence, years spent in formal education, health insurance coverage, smoking status and alcohol use. Abbreviations as elsewhere defined.

In contrast, there was a more discernible J-shaped relationship between (unadjusted and adjusted) mean HbA1c and each obesity index among females (**Figures 2b, d and f**). At higher values of BMI (>21 kg/m^2^), WtHR (>0.48) and WC (>78 cm), the adjusted mean HbA1c was higher in female PWOH than female PWH. Conversely, below these thresholds, adjusted mean HbA1c was higher in PWH than PWOH with decreasing BMI, WtHR or WC.

### Association between prevalent diabetes, HIV and obesity indices

Diabetes was more prevalent among PWOH (15.9%) than PWH (9.0%; p<0.001), overall (**Table 1**). In both unadjusted (**Supplementary Table 1**) and adjusted (**Table 2**) sex stratified analyses, however, there were no significant differences in diabetes prevalence between male PWH and PWOH, both overall and by obesity categories. This contrasted sharply with females among whom PWOH [15.7% (14.4, 17.0)] had higher diabetes prevalence than PWH [10.5% (8.3, 12.8); p<0.001], corresponding to a prevalence difference of -5.2% (-7.8, -2.6) (**Table 3**). These HIV-related differences in diabetes prevalence among females were more pronounced with increasing obesity. For example, diabetes prevalence at BMI ≥30kg/m^2^ was 12.8% (9.1, 16.5) among female PWH versus 22.0% (19.9, 24.0) among female PWOH (p<0.001). That is, females with HIV and obesity had half the relative prevalence of diabetes than female PWOH [adjusted prevalence ratio (adjPR) 0.58 (0.41, 0.76); p<0.001] (**Table 3**). These observation were largely similar in the unadjusted analyses (**Supplementary Table 2**).

### Sociodemographic and behavioral determinants of HbA1c and prevalent diabetes

Among males, age, smoking status and household wealth index were significantly associated with mean HbA1c and diabetes prevalence (**Supplementary Table 3**). Whereas age and household wealth index were positively correlated with diabetes prevalence, current smoking (versus none) was inversely associated. On the other hand, only age and alcohol use were significant determinants of diabetes prevalence among females in addition to HIV serostatus. Current drinking status (versus non-drinking) was negatively correlated with diabetes prevalence.

### Sensitivity analyses

Female PWOH [19.9% (17.8, 22.0)] had greater diabetes prevalence than female PWH [12.5% (8.9, 16.0); p<0.011] in MFP analyses using a higher inclusion age (≥40 vs. ≥20 years old) and a diabetes definition based solely on HbA1c (**Table 3**). That is, the relative prevalence of diabetes among female PWH ≥40 years old was nearly two-thirds that of female PWOH [adjPR 0.62 (0.43, 0.82); p<0.001] (**Table 3**). However, diabetes prevalence was similar between male (≥40 years old) PWOH [13.3% (10.9, 15.8)] and male PWH [8.0% (3.6, 12.5); p=0.414]. PS matching (**Supplementary Figure 1**) produced results that were consistent with both MFP-based primary and secondary analyses (**Table 3**). For example, the adjPR for diabetes among females was 0.67 (0.51, 0.82) for the primary MFP model, 0.62 (0.43, 0.82) for secondary MFP model and 0.63 (0.45, 0.81) for the secondary PS matched model.

## Discussion

In a large national sample of South African adults we found no HIV-related differences in prevalent diabetes risk or HbA1c distribution among males. In contrast, female PWH had a lower prevalence of diabetes and lower mean HbA1c than peers without HIV. Notably, these differences were greatest among those living with obesity. Our data were cross-sectional and warrant cautious interpretation. They nonetheless raise important questions. Whether PWH in SSA have increased risk of diabetes remains to be ascertained, at least in the South African context. Among women living with HIV and obesity in particular, the comparatively reduced risk of diabetes suggests possible protective adaptations, physiological or otherwise. Future work in South Africa and other HIV-endemic SSA settings should be directed towards better mechanistic understanding of sex-specific adipose tissue biology in HIV as well as map its implications for diabetes risk, clinical phenotypes, management and prognosis.

Excess fat accumulation in visceral depots is a major cause of diabetes^24, 26, 41^ via worsening of insulin sensitivity^24, 27^ and/or loss of β-cell function and islet mass.^26, 27^ Current understanding, albeit gained from studies^29, 30, 42^ in HICs, is that HIV/ART and obesity synergistically drive the excess diabetes risk in both male and female PWH compared to PWOH.^9, 10, 11^ HIV/ART are thought to induce deleterious qualitative, quantitative and distributive changes in adipose tissue.^42, 43^ Mechanistically, HIV infects adipose tissue resident CD4+ T cells and macrophages.^42^ ART causes mitochondrial toxicity, oxidative stress and altered gene expression in adipocytes.^44^ This is in addition to causing weight gain. The resulting and frequently excessive by adipose tissue is characterized by, among other disorders, increased insulin resistance.^42^ Our findings appear, however, to contradict this paradigm. Male PWH had similar diabetes risk to male PWOH at all levels of adiposity, whereas diabetes risk in female PWH was reduced with obesity but accentuated with leanness.

There are a number of potential explanations for our findings. First, BMI, WC and WtHR are inadequate indices of fat accumulation. BMI poorly discriminates between lean and fat mass. WC and WtHR, on the other hand, more accurately measure central obesity although they cannot distinguish between abdominal subcutaneous (SAT) and visceral adipose tissue (VAT) depots. Neither can they measure ectopic sites like hepatic, pancreatic, and omental adipose tissue.^43^ Different fat depots and ectopic sites have different dysmetabolic potential.^43^ Relatedly, HIV/ART are known to influence the partitioning and distribution of adipose tissue.^43, 45, 46^ Thus, similar BMI, WC or WtHR between PWH and PWOH may not reflect similar diabetes risk as the underlying adipose tissue composition and distribution can vary substantially. In South Africa, for example, Goedecke *et al*., (2013)^45^ used dual energy X-ray absorptiometry (DXA) and found that despite similar BMI, ART-experienced female PWH had greater VAT and lesser leg fat than ART-naive female PWH. Our hypothesis is, therefore, that increasing BMI, WC and WtHR in our cohort corresponded to increasing fat accumulation albeit in less diabetogenic depots and/or ectopic sites in female but not male PWH compared to PWOH.

This hypothesis may assume greater relevance in light of growing evidence associating newer generation integrase inhibitors (INSTI) with weight gain and obesity, and the subsequent concerns about diabetes risk.^47, 48, 49^ Of note, INST-related weight gain disproportionally affects women and those of Black ethnicity. Their uptake is also increasing across SSA since their recommendation by WHO as preferred first line ART. On the other hand, the literature from SSA on diabetes risk associated with INSTI-related weight gain is presently very sparse and contradictory.^47, 48, 49^ INSTI became publicly available in South Africa since 2019 whereas data collection for our study took place in 2016. It, therefore, remains to be determined how excess adipose tissue gained from INSTI use is partitioned and impacts diabetes risk.

It also remains to be established how, if, HIV/adiposity interactions shape clinical phenotype(s) of diabetes in SSA. The presence of diabetes phenotypes in the general population in SSA that may not fit the conventional classification of (type II) diabetes has already been noted.^22, 50^ For example, the majority of patients with diabetes in the general population in SSA are young, lean, and have β-cell secretory dysfunction more than insulin resistance compared to those in HICs.^22, 50^ Our finding of worse glucose profiles in female PWH compared to female PWOH at lower adiposity levels may speak to this pathophysiological heterogeneity and, in turn, the phenotypic diversity of diabetes in SSA. Exploration and subsequent identification of distinct HIV-related diabetes phenotypes, if they exist, is therefore critical to informing optimal therapeutic approaches and preventive strategies among SSÀs 26 million PWH.^51^ Future work in South Africa should combine imaging-based assessment of adipose tissue depots and ectopic sites; detailed assessments of glycemic and insulin indices using, for example, 2 hour multiple-sampled oral glucose oral testing; together with adipose tissue sampling and molecular characterization.

### Strengths and limitations

Our participants were drawn from across South Africàs racial/ethnic, demographic, socioeconomic and geospatial groups. However, we did not apply sampling weights to our analyses and thus our results are not population-level estimates. Similarly, the cross-sectional design forestalls causal inferences and warrants caution in interpretation of these results. While the large sample size permitted examination of important subgroups, our lack of data on CD4+ counts, ART history and other HIV clinical measures was another limitation. Persons LWH on ART versus those not on ART, for example, are likely to differ in terms of cardiometabolic and inflammation profiles in ways that may impact our findings. We also cannot not determine the contribution, if any, of primary care access to the observed effects. Likewise, our tools to assess body size as a marker of adiposity were comparatively blunt.

Our definition of diabetes is another major limitation. Oral glucose tolerance testing is the diagnostic gold standard. Data from South Africa^52^ and elsewhere^53^ show that the HbA1c threshold of 6.5% has low sensitivity compared to lower thresholds, e.g., 5.8%, for diagnosing diabetes among PWH. On the other hand, the limitation of single HbA1c measurement in community-based diabetes screening are highlighted by the observation that a screening threshold of HbA1c of >16.6% is needed to ensure a 90% positive predictive value.^54^

## Conclusion

In a large sample of people with and without HIV in South Africa, we found glycemic profiles were similar between people with and without HIV. In fact, women living with HIV and obesity in particular had comparatively lower risk of diabetes than women living with obesity without HIV. These data call into question whether PWH in the South African context are at increased risk of diabetes. The mechanisms, physiological or otherwise, conferring this potential protection from diabetes, particularly at greater obesity, must be identified. Future work in South Africa and other HIV-endemic SSA settings should be directed towards better mechanistic understanding of sex-specific adipose tissue biology in HIV, especially with increasing INSTI use and the associated higher odds of weight gain and obesity.

## Data Availability

All the data reported in this analysis are freely accessible from the Demographic Health Survey (DHS) Program. https://dhsprogram.com/

## Author Contributions

IMM, ACC, MJS – conceptualization; IMM - data curation; IMM - formal analysis; IMM, ACC, NT, JHG, RS, JMG, NABN, MJN, MJS – investigation; IMM, ACC, NT, JHG, RS, JMG, NABN, MJN, MJS – methodology; IMM - project administration; validation, IMM - visualization, IMM -writing – original draft; and IMM, ACC, NT, JHG, RS, JMG, NABN, MJN, MJS - writing – review & editing.

All authors had full access to all the data in the study and had final responsibility for the decision to submit for publication.

---

**Supplementary Figure 1.**
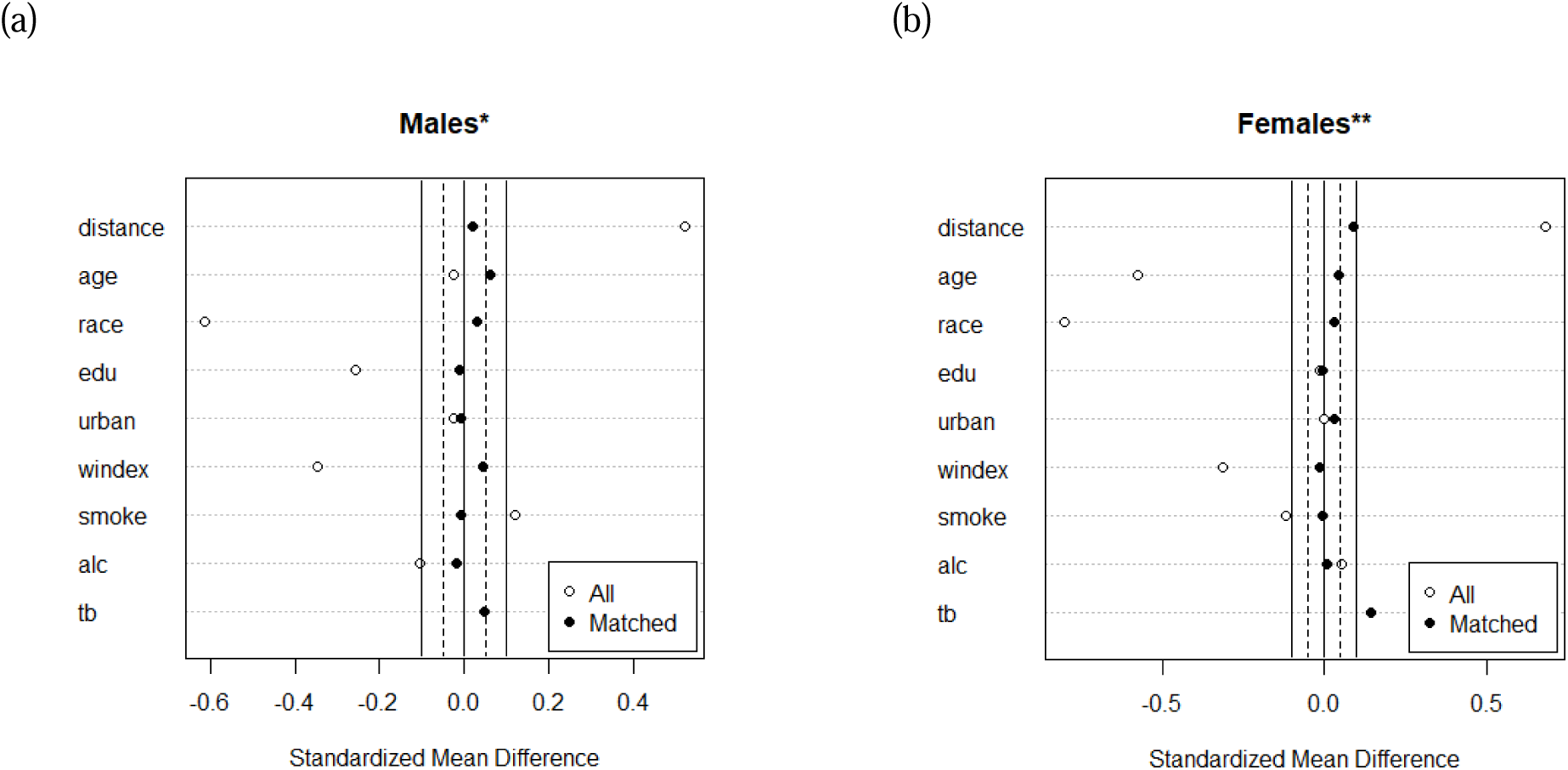
Covariate balance before and after propensity score matching of (a) men and (b) women (≥20 years old) by HIV serostatus for sensitivity analysis: unweighted South Africa Demographic Health Survey, 2016. * Among men, there were 1,634 PWOH and 312 PWH before matching; and 312 PWOH and 312 PWH after matching. ** Among women, there were 2,584 PWOH and 901 PWH before matching; and 901 PWOH and 901 PWH after matching.

**Supplementary Figure 2.**
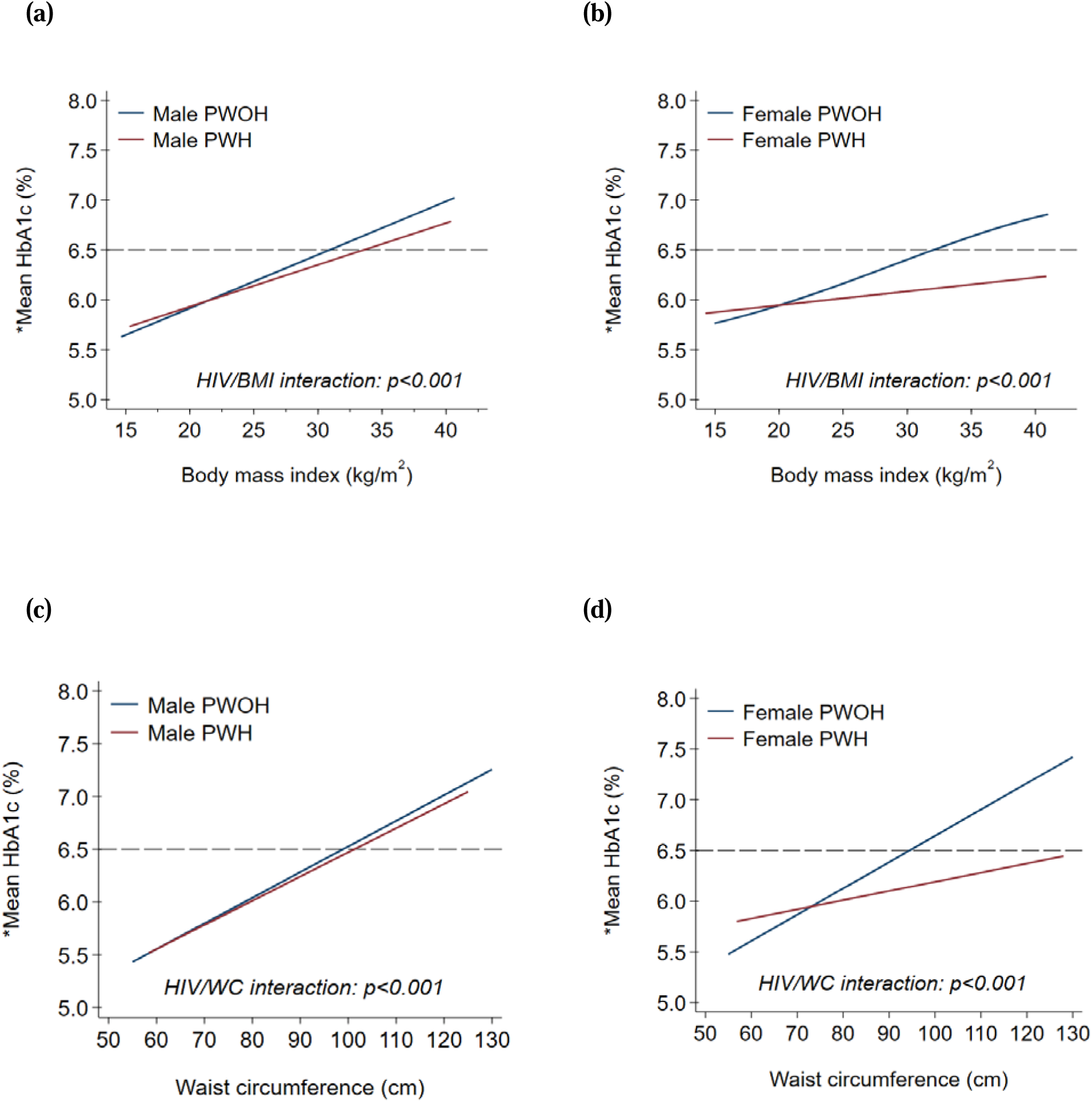
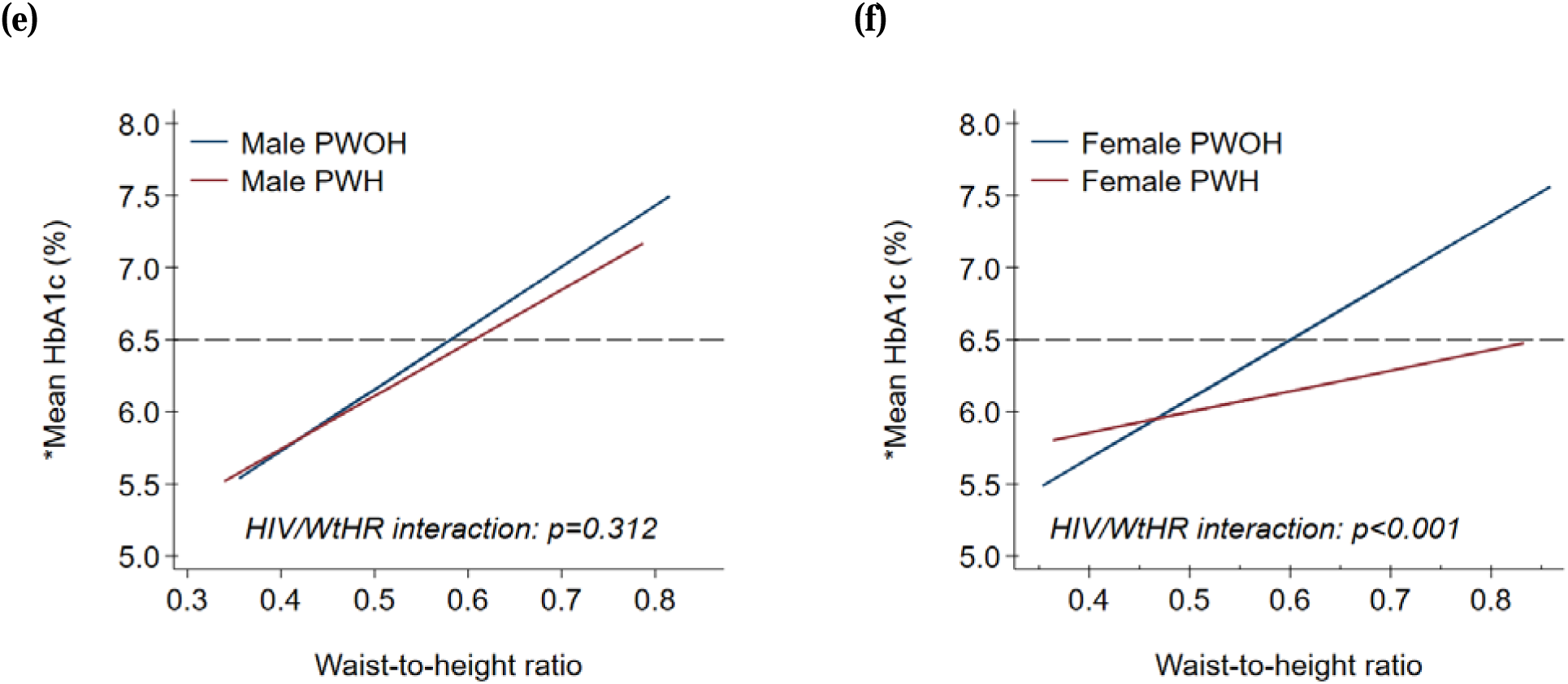
Mean glycated (HbA1c) according to waist circumference, body mass index and waist-to-height ratio by HIV serostatus and sex among adults aged ≥20 years old: unweighted South Africa Demographic Health Survey, 2016. * Estimated from univariable fractional polynomial model of specific anthropometric index without adjustment. Abbreviations as elsewhere defined.

