## Supplementary material for "Associations of HIV and prevalent type 2 diabetes mellitus in the context of obesity in South Africa": Tables 2, 3 & 4; Suppl Tables 2 & 3

**Table 2. Adjusted prevalence, prevalence difference and prevalence ratio of type 2 diabetes mellitus according to waist circumference, waist-to-height ratio and body mass index categories by HIV serostatus for males ≥20 years old: unweighted South Africa Demographic Health Survey, 2016**

| **Adiposity category** | **Adjusted^α^ estimate (95%CI); *Men*** | | | | | |
| --- | --- | --- | --- | --- | --- | --- |
|  | **Prevalence** |  | **Prevalence** | **P value** | **Prevalence** | **P value** |
|  | **PWOH** | **PWH** | **difference**^β^ |  | **ratio**^β^ |  |
| Overall | 9.8 (8.5, 11.1) | 11.3 (7.1, 15.5) | 1.5 (-2.9, 5.9) | 0.660 | 1.15 (0.69, 1.61) | 0.740 |
| *Waist circumference* | | | | | | |
| Normal | 6.3 (4.8, 7.8) | 5.3 (2.3, 8.3) | -1.0 (-4.3, 2.3) | 0.547 | 0.84 (0.33, 1.35) | 0.774 |
| Elevated | 15.7 (12.6, 18.7) | 21.4 (11.7, 31.0) | 5.7 (-4.4, 15.8) | 0.269 | 1.37 (0.69, 2.4) | 0.516 |
| *Waist-to-height ratio* | | | | | | |
| Normal | 4.8 (3.1, 6.5) | 4.4 (1.2, 7.7) | -0.4 (-4.1, 3.3) | 0.839 | 0.92 (0.17, 1.67) | 0.899 |
| Elevated | 13.2 (11.0, 15.4) | 13.7 (8.0, 19.4) | 0.5 (-5.7, 6.6) | 0.868 | 1.04 (0.57, 1.50) | 0.932 |
| *Body mass index* | | | | | | |
| Underweight | 2.6 (-0.3, 5.6) | 5.6 (-2.3, 13.5) | 2.9 (-5.4, 11.3) | 0.689 | 2.11 (-1.66, 5.89) | 0.515 |
| Normal | 7.4 (5.4, 9.3) | 5.2 (1.7, 8.8) | -2.1 (-6.2, 1.9) | 0.295 | 0.71 (0.19, 1.22) | 0.284 |
| Overweight | 11.6 (9.0, 14.2) | 11.6 (4.4, 18.8) | 0.0 (-7.6, 7.6) | 0.998 | 1.00 (0.34, 1.65) | 0.714 |
| Obese | 15.7 (12.0, 19.5) | 26.8 (8.4, 45.2) | 11.1 (-7.7, 29.9) | 0.249 | 1.70 (0.46, 2.95) | 0.769 |

^α^ Variables and their transformation for which adjustment was made using multivariable fractional polynomial models are outlined in Supplementary Table 1.

^β^ Persons LWOH are the reference group.

Elevated waist circumference if ≥94cm (for males). Elevated waist-to-height ratio if ≥0.5. Body mass index categories: underweight: <18.5 kg/m^2^; normal 18.5-24.9 kg/m^2^; overweight 25-29.9 kg/m^2^; obese ≥30 kg/m^2^.

Abbreviations as elsewhere defined.

**Table 3. Adjusted prevalence, prevalence difference and prevalence ratio of type 2 diabetes mellitus according to waist circumference, waist-to-height ratio and body mass index categories by HIV serostatus for females ≥20 years old: unweighted South Africa Demographic Health Survey, 2016**

| **Adiposity category** | **Adjusted^α^ estimate (95%CI); *Women*** | | | | | |
| --- | --- | --- | --- | --- | --- | --- |
|  | **Prevalence** |  | **Prevalence** | **P value** | **Prevalence** | **P value** |
|  | **PWOH** | **PWH** | **difference**^β^ |  | **ratio**^β^ |  |
| Overall | 15.7 (14.4, 17.0) | 10.5 (8.3, 12.8) | -5.2 (-7.8, -2.6) | <0.001 | 0.67 (0.51, 0.82) | <0.001 |
| *Waist circumference* | | | | | | |
| Normal | 5.1 (3.3, 7.0) | 6.2 (2.9, 9.4) | 1.0 (-2.7, 4.8) | 0.595 | 1.20 (0.43, 1.97) | 0.551 |
| Elevated | 18.5 (17.0, 20.1) | 11.7 (9.0, 14.4) | -6.9 (-10.0, -3.7) | <0.001 | 0.63 (0.48, 0.78) | <0.001 |
| *Waist-to-height ratio* | | | | | | |
| Normal | 6.4 (4.1, 8.7) | 8.3 (4.5, 12.1) | 1.9 (-2.3, 6.3) | 0.241 | 1.30 (0.55, 2.05) | 0.241 |
| Elevated | 19.5 (18.0, 21.1) | 12.1 (9.3, 14.8) | -7.5 (-10.7, -4.3) | <0.001 | 0.62 (0.46, 0.77) | <0.001 |
| *Body mass index* | | | | | | |
| Underweight | 4.6 (-1.4, 10.5) | 7.8 (-2.9, 18.5) | 3.3 (-8.9, 15.4) | 0.602 | 1.72 (-1.53, 4.96) | 0.756 |
| Normal | 7.2 (5.1, 9.3) | 7.1 (3.4, 10.8) | -0.1 (-4.4, 4.1) | 0.956 | 0.98 (0.39, 1.57) | 0.944 |
| Overweight | 11.6 (9.4, 13.8) | 9.3 (5.3, 13.2) | -2.3 (-6.8, 2.2) | 0.312 | 0.80 (0.43, 1.17) | 0.585 |
| Obese | 22.0 (19.9, 24.0) | 12.8 (9.1, 16.5) | -9.2 (-13.4, -4.9) | <0.001 | 0.58 (0.41, 0.76) | <0.001 |

^α^ Variables and their transformation for which adjustment was made using multivariable fractional polynomial models are outlined in Supplementary Table 1.

^β^ PWOH uninfected are the reference group.

Elevated waist circumference if ≥80cm (for females). Elevated waist-to-height ratio if ≥0.5. Body mass index categories: underweight: <18.5 kg/m^2^; normal 18.5-24.9 kg/m^2^; overweight 25-29.9 kg/m^2^; obese ≥30 kg/m^2^.

Abbreviations as elsewhere defined.

**Table 4.** Sensitivity analysis of adjusted prevalence, prevalence difference and prevalence ratio of type 2 diabetes mellitus according to HIV serostatus and sex: unweighted South Africa Demographic Health Survey, 2016

| **Analytic Approach*** | **Adjusted^α^ estimate (95%CI)** | | | | | |
| --- | --- | --- | --- | --- | --- | --- |
|  | **Prevalence** |  | **Prevalence** | **P value** | **Prevalence** | **P value** |
|  | **PWOH** | **PWH** | **difference**^β^ |  | **ratio**^β^ |  |
| ***Men: Diabetes*** |  |  |  |  |  |  |
| Primary, *MFP model* | 9.8 (8.5, 11.1) | 11.3 (7.1, 15.5) | 1.5 (-2.9, 5.9) | 0.660 | 1.15 (0.69, 1.61) | 0.740 |
| Secondary, *MFP model* | 13.3 (10.9, 15.8) | 8.0 (3.6, 12.5) | -5.1 (-10.4, 2,1) | 0.414 | 0.60 (0.34, 1.08) | 0.089 |
| Secondary, *PS matching* | 8.3 (5.3, 11.4) | 7.7 (4.7, 10.7) | -0.6 (-0.4, 3.6) | 0.768 | 0.92 (0.43, 1.41) | 0.768 |
| ***Women: Diabetes*** | | | | | | |
| Primary, *MFP model* | 15.7 (14.4, 17.0) | 10.5 (8.3, 12.8) | -5.2 (-7.8, -2.6) | <0.001 | 0.67 (0.51, 0.82) | <0.001 |
| Secondary, *MFP model* | 19.9 (17.8, 22.0) | 12.5 (8.9, 16.0) | -7.5 (-11.7, -3.3) | 0.003 | 0.62 (0.43, 0.82) | <0.001 |
| Secondary, *PS matching* | 12.8 (10.6, 14.9) | 8.1 (6.3, 9.9) | -4.7 (-7.5, -18.5) | 0.001 | 0.63 (0.45, 0.81) | 0.001 |

^α^ Adjusted for age, race, urban/rural residence, household wealth index, smoking and alcohol status, and past TB treatment.

^β^ PWOH are the reference group.

* Primary, *MFP model*: diabetes = HbA1c ≥6.5% and/or current use of oral hypoglycemic medicines and/or insulin among adults ≥20 years old; using multivariable fractional polynomial generalized linear models (GLM).

Secondary, *MFP model*: diabetes = HbA1c ≥6.5% and taking neither oral hypoglycemic medicines nor insulin among adults ≥40 years old; using multivariable fractional polynomial GLM.

Secondary, *PS matching*: diabetes = HbA1c ≥6.5% and/or current use of oral hypoglycemic medicines and/or insulin among adults ≥20 years old; using propensity score matched GLM.

**Supplementary Table 1. Unadjusted prevalence of type 2 diabetes mellitus according to waist circumference, waist-to-height ratio and body mass index categories by HIV serostatus and sex males aged ≥20 years old: unweighted South Africa Demographic Health Survey, 2016**

| **Adiposity category** | **Unadjusted estimate (95%CI); *Males*** | | | | | |
| --- | --- | --- | --- | --- | --- | --- |
|  | **Prevalence** |  | **Prevalence** | **P value** | **Prevalence** | **P value** |
|  | **PWOH** | **PWH** | **difference**^α^ |  | **ratio**^α^ |  |
| Overall | 10.0 (8.6, 11.3) | 9.2 (5.9, 12.5) | -0.8 (-4.4, 2.7) | 0.651 | 0.92 (0.56, 1.27) | 0.117 |
| *Waist circumference* | | | | | | |
| Normal | 5.1 (3.9, 6.3) | 4.4 (2.0, 6.8) | -0.7 (-3.4, 2.0) | 0.618 | 0.86 (0.34, 1.38) | 0.635 |
| Elevated | 26.8 (22.4, 31.1) | 25.5 (13.5, 37.5) | -1.3 (-14.0, 11.5) | 0.844 | 0.95 (0.48, 1.42) | 0.847 |
| *Waist-to-height ratio* | | | | | | |
| Normal | 3.2 (2.1, 4.3) | 3.4 (0.9, 5.9) | 0.2 (-2.5, 2.9) | 0.896 | 1.06 (0.20, 1.91) | 0.895 |
| Elevated | 20.2 (17.2, 23.20 | 15.5 (8.9, 22.1) | -4.7 (-11.9, 2.6) | 0.204 | 0.77 (0.42, 1.11) | 0.251 |
| *Body mass index* |  |  |  |  |  |  |
| Underweight | 2.3 (-0.3, 4.9) | 5.3 (-1.8, 12.4) | 2.9 (-4.6, 10.5) | 0.443 | 2.28 (-1.72, 6.28) | 0.358 |
| Normal | 5.5 (4.0, 6.9) | 4.1 (1.3, 7.0) | -1.3 (-4.5, 1.8) | 0.408 | 0.76 (0.20, 1.31) | 0.452 |
| Overweight | 15.6 (12.0, 19.3) | 12.5 (4.4, 20.6) | -3.1 (-12.0, 57.6) | 0.490 | 0.80 (0.25, 1.35) | 0.526 |
| Obese | 26.5 (20.8, 32.2) | 26.1 (8.1, 44.0) | -0.4 (-0.19.3, 18.4) | 0.964 | 0.98 (0.27, 1.69) | 0.964 |

^α^ PWOH are the reference group.

Elevated waist circumference if ≥94cm (for males). Elevated waist-to-height ratio if ≥0.5. Body mass index categories: underweight: <18.5 kg/m^2^; normal 18.5-24.9 kg/m^2^; overweight 25-29.9 kg/m^2^; obese ≥30 kg/m^2^.

Abbreviations as elsewhere defined.

**Supplementary Table 2. Unadjusted prevalence of type 2 diabetes mellitus according to waist circumference, waist-to-height ratio and body mass index categories by HIV serostatus and sex among females aged ≥20 years old: unweighted South Africa Demographic Health Survey, 2016**

| **Adiposity category** | **Unadjusted estimate (95%CI); *Females*** | | | | | |
| --- | --- | --- | --- | --- | --- | --- |
|  | **Prevalence** |  | **Prevalence** | **P value** | **Prevalence** | **P value** |
|  | **PWOH** | **PWH** | **difference**^α^ |  | **ratio**^α^ |  |
| Overall | 16.7 (15.3, 18.1) | 8.5 (6.7, 10.4) | -8.2 (-10.5, -5.9) | <0.001 | 0.51 (0.39, 0.63) | <0.001 |
| *Waist circumference* | | | | | | |
| Normal | 3.8 (2.4, 5.1) | 4.2 (2.0, 6.4) | 0.4 (-2.2, 3.1) | 0.318 | 1.11 (0.39, 1.83) | 0.746 |
| Elevated | 22.1 (20.2, 23.9) | 10.3 (7.9, 12.8) | -11.7 (-14.8, -8.7) | <0.001 | 0.47 (0.36, 0.59) | <0.001 |
| *Waist-to-height ratio* | | | | | | |
| Normal | 3.7 (2.2, 5.2) | 5.1 (2.6, 7.6) | 1.4 (-1.5, 4.30 | 0.953 | 1.38 (0.51, 2.25) | 0.314 |
| Elevated | 21.4 (19.6, 23.3) | 9.7 (7.4, 12.1) | -11.7 (-14.7, -8.7) | <0.001 | 0.45 (0.34, 0.57) | <0.001 |
| *Body mass index* |  |  |  |  |  |  |
| Underweight | 3.7 (-1.3, 8.7) | 6.7 (-2.3, 15.6) | 2.9 (-7.3, 13.2) | 0.567 | 1.80 (-1.64, 5.23) | 0.548 |
| Normal | 6.4 (4.5, 8.2) | 4.8 (2.3, 7.4) | -1.5 (-4.7, 1.7) | 0.348 | 0.76 (0.29, 1.22) | 0.377 |
| Overweight | 11.9 (9.5, 14.2) | 6.9 (3.9, 9.9) | -5.0 (-8.8, -1.2) | 0.009 | 0.58 (0.30, 0.86) | 0.025 |
| Obese | 26.7 (24.2, 29.3) | 11.6 (8.2, 15.1) | -15.1 (-19.4, -10.8) | <0.001 | 0.44 (0.31, 0.57) | <0.001 |

^α^ PWOH are the reference group.

Elevated waist circumference if ≥80cm (for females). Elevated waist-to-height ratio if ≥0.5. Body mass index categories: underweight: <18.5 kg/m^2^; normal 18.5-24.9 kg/m^2^; overweight 25-29.9 kg/m^2^; obese ≥30 kg/m^2^.

Abbreviations as elsewhere defined.

**Supplementary** **Table 3. Parameters for multivariate fractional polynomial models estimating adjusted prevalence, prevalence difference and prevalence ratio for type 2 diabetes mellitus by anthropometric categories among adults aged ≥20 years old: South Africa Demographic Health Survey, 2016, unweighted**

| **Variable (X)** | **Men** | | | | **Women** | | |
| --- | --- | --- | --- | --- | --- | --- | --- |
|  | **Transformation** | **β** | **SE** | **Transformation** | | **β** | **SE** |
| ***Waist circumference*** | | | | | | | |
| HIV status | - | - | - | X | | -0.383 | 0.129 |
| WC, categorical | X | 0.976 | 0.164 | X | | 1.115 | 0.166 |
| Age | (X/10)^3^ - 73.146 | 0.054 | 0.008 | ln(X/10) – 1.484 | | 3.317 | 0.346 |
| Age | [(X/10)^3^ x ln((X/10)] - 104.659 | -0.024 | 0.004 | (X/10)^3^ – 86.013 | | -0.004 | 0.006 |
| Alcohol use | - | - | - | X | | -0.734 | 0.169 |
| Smoking status | X | -0.351 | 0.174 | - | | - | - |
| Wealth index | X + 0.216 | 0.017 | 0.008 | - | | - | - |
| ***Waist-to-height ratio*** | | | | | | | |
| HIV status | - | - | - | X | | -0.379 | 0.129 |
| WtHR, categorical | X | 1.054 | 0.200 | X | | 0.965 | 0.168 |
| Age | (X/10)^3^ -73.064 | 0.053 | 0.009 | ln(X/10) – 1.484 | | 3.400 | 0.345 |
| Age | [(X/10)^3^ x ln((X/10)] -104.514 | -0.024 | 0.004 | (X/10)^3^ – 85.768 | | -0.003 | 0.006 |
| Alcohol use | - | - | - | X | | -0.738 | 0.169 |
| Smoking status | X | -0.334 | 0.176 | - | | - | - |
| Wealth index | X + 0.207 | 0.024 | 0.008 | - | | - | - |
| ***Body mass index*** | | | | | | | |
| HIV status | - | - | - | X | | -0.376 | 0.131 |
| BMI, categorical | X - 1.396 | 0.449 | 0.089 | X - 2.126 | | 0.526 | 0.064 |
| Age | (X/10)^3^ - 72.492 | 0.058 | 0.009 | ln(X/10) – 1.485 | | 3.216 | 0.346 |
| Age | [(X/10)^3^ x ln((X/10)] - 103.506 | -0.027 | 0.004 | (X/10)^3^ – 86.009 | | -0.003 | 0.006 |
| Alcohol use | - | - | - | X | | -0.644 | 0.171 |
| Smoking status | X | -0.299 | 0.177 |  | |  |  |
| Wealth index | X + 0.159 | 0.020 | 0.008 | - | | - | - |

Models were sex-stratified and initially included age, race, area of residence, household wealth index, smoking and alcohol drinking status, and history of TB drug treatment for TB use as covariates. Only covariates that were statistically significant at p-value <0.05 were retained in final models and are reported in the table included their transformations.

Age is divided by 10 before transformation to improve the scaling of the regression (β) coefficients.

WC = waist circumference; WtHR = waist-to-height ratio; BMI = body mass index; In = natural log transformation.
